# Kinematic analysis of upper body movements in adolescent with mild idiopathic scoliosis: a preliminary study

**DOI:** 10.1101/2021.06.09.21258605

**Authors:** Lucas Struber, Aurélien Courvoisier, Jacques Griffet, Olivier Daniel, Alexandre Moreau-Gaudry, Philippe Cinquin, Vincent Nougier

## Abstract

Analysis of kinematic and postural data of adolescent idiopathic scoliosis (AIS) patients seems relevant for a better understanding of biomechanical aspects involved in AIS and its etiopathogenesis. The present project aimed at investigating kinematic differences and asymmetries in early AIS in a static task and in uniplanar trunk movements (rotations, lateral bending and forward bending). Trunk kinematics and posture were assessed using a 3D motion analysis system and a force plate. Fifteen healthy girls, fifteen AIS girls with left lumbar main curve and seventeen AIS girls with right thoracic main curve were compared. Statistical analyses were performed to investigate presumed differences between the three groups. This study showed kinematic and postural differences between mild AIS patients and controls such as static imbalance, a reduced range of motion in the frontal plane and a different kinematic strategy in lateral bending. These differences mainly occurred in the same direction whatever the type of scoliosis, and suggested that AIS patients behave similarly from a dynamic point of view.

## Introduction

Adolescent idiopathic scoliosis (AIS) is a structural, lateral, rotated curvature of the spine that affects 1-3 % of children and arises around puberty [1]. Relative orientations of trunk segments have been shown to exhibit differences related both to the severity [2] and type of scoliosis [3]. AIS has also been associated with postural instability [4,5], sensory integration deficits [5,6] and alteration of gait pattern combined to a reduced range of motion in the upper body and lower extremities [7,8]. However, observed differences are not consistent, relatively small in comparison to a healthy population and most of these studies were limited to one type of curve or mixed up all curve types and Cobb angles. These results suggested that analysis of kinematic and postural differences between healthy subjects and AIS patients is clinically relevant for a better understanding of AIS biomechanics. However, a better methodology is necessary to come up with a more complete description.

The main objective of this preliminary study was to detect on mild scoliosis kinematic and postural differences in the different planes of motion of simple uniplanar upper body movements. We focused on mild scoliosis in order to detect singularities that are already present at an early stage of the disease and which then could be potential markers of aggravation. We hypothesized that AIS patients exhibited 1) a static postural imbalance, 2) a smaller upper body range of motion and 3) left and right asymmetrical motion depending on the side of the deformation.

## Materials and methods

### Participants

47 adolescents aged between 9 and 16 years voluntarily participated in the study (Table 1). 15 healthy girls composed the control group (HS), 15 AIS girls with left lumbar main curve composed the AIS-LL group and 17 AIS girls with right thoracic main curve composed the AIS-RT group, respectively type 1 and 5 according to Lenke classification [9]. For the two AIS groups, radiological data, Cobb angle [10], and Risser sign [11] were evaluated from standard standing anteroposterior radiography. In the AIS groups, the patients included in the study were not braced or were included in the study before bracing. The local ethics committee (Comité de Protection des Personnes Sud-Est V) approved this research (Ref. CPP 14-CHUG-14), all methods were performed in accordance with the relevant guidelines and regulations and written informed consents were obtained from all the subjects and their parents.

**Table 1.**
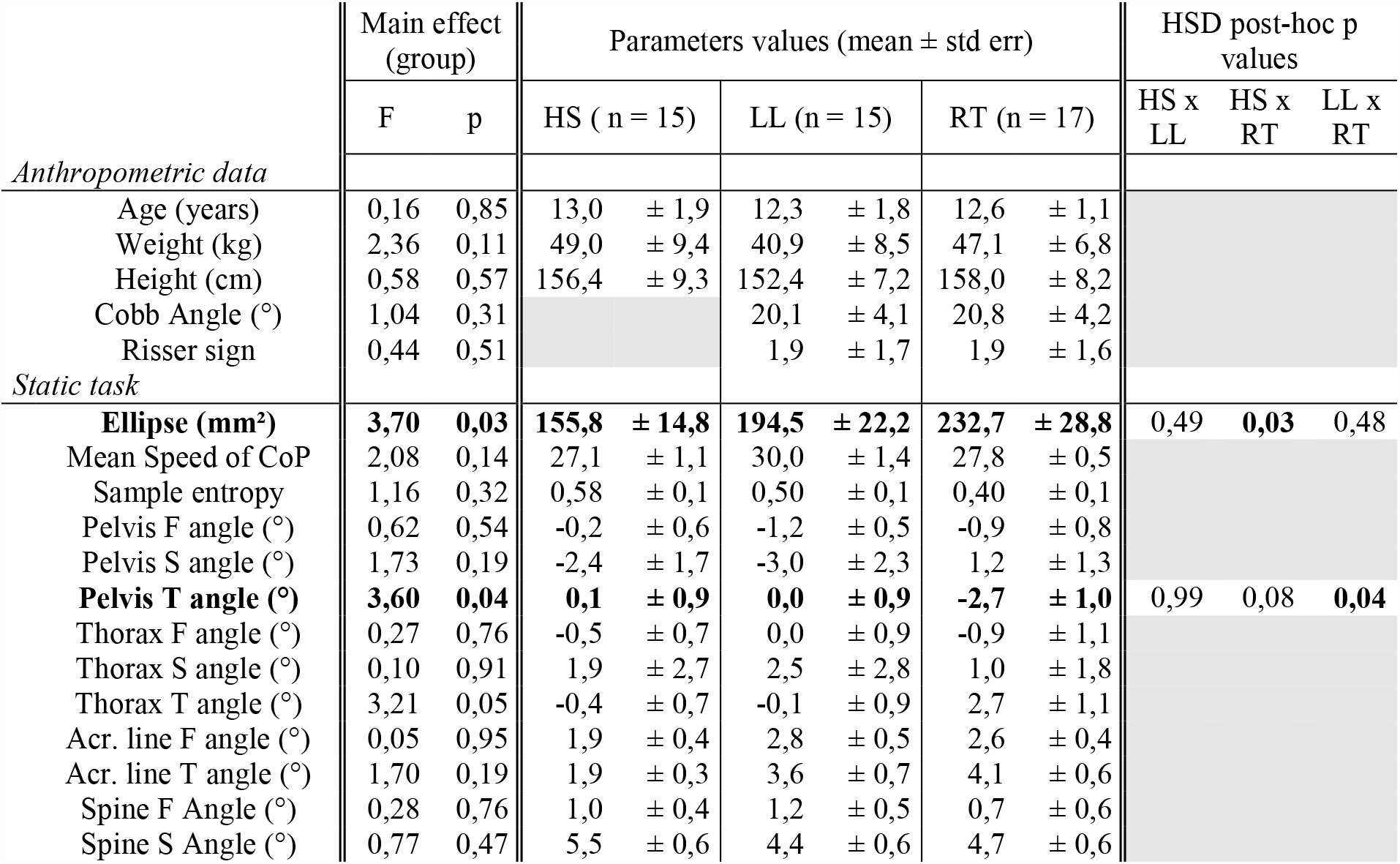
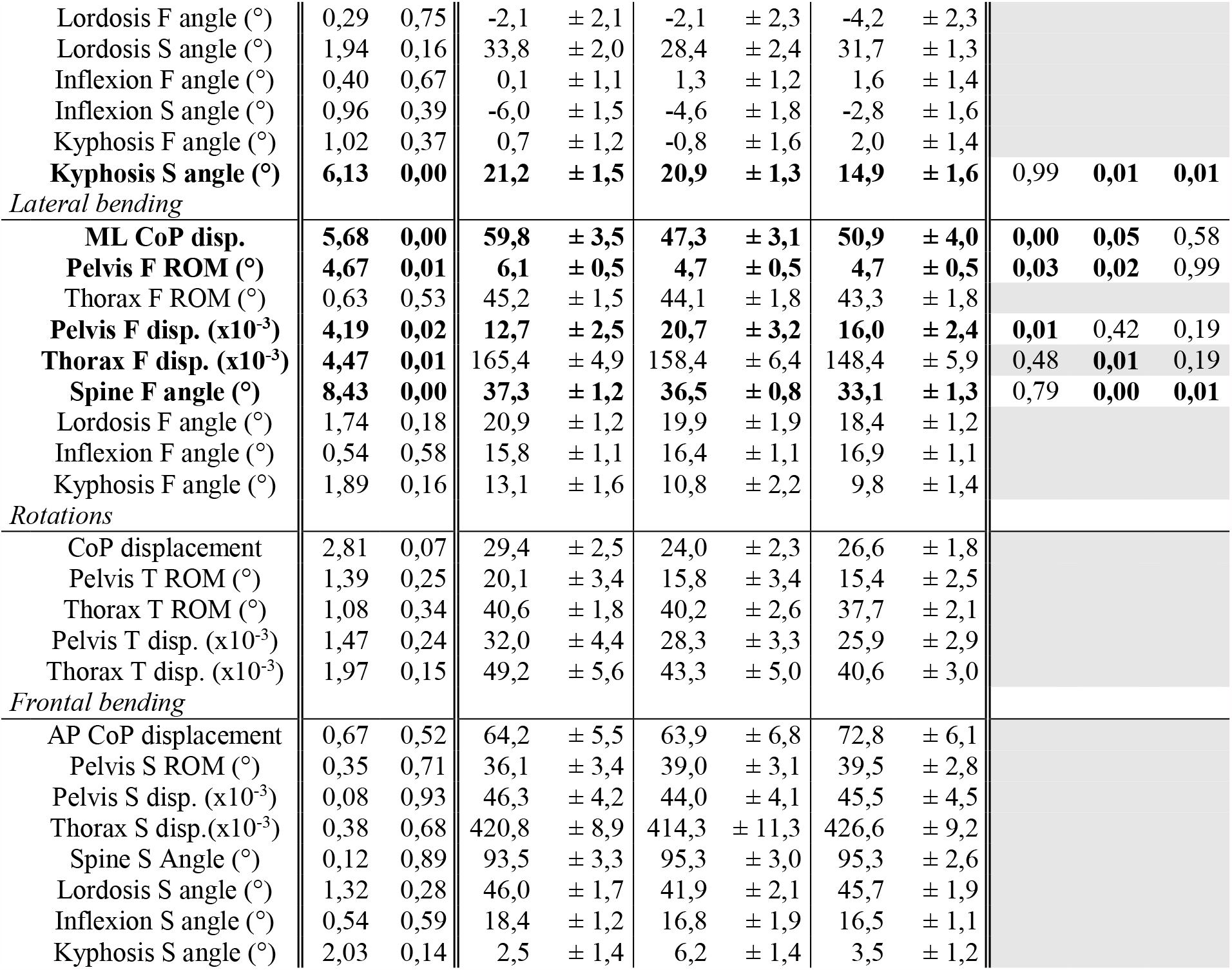
Summary of results. In lateral bending and rotation tasks, values from right and left movements were averages since no effect of side was observed. F: Frontal, S: Sagittal, T: Transverse.

### Experimental tasks

Participants started the experiment performing a static task. They stood upright and barefoot, arms along the body, heels 5 cm apart with internal edges of the feet making a 30° angle. They were instructed to focus on a cross target located at eyes level and to remain in a stable relaxed posture for 30 sec. 4 trials were performed. Then, starting upright, participants performed three dynamic tasks involving the upper body: 1) forward maximal bending with arms along the body, 2) right and left maximal lateral bending with arms along the body, and 3) right and left maximal axial rotations with arms crossed on the chest. Each task was repeated 4 times.

### Data recording

A force plate recorded center of feet pressure (CoP) displacements (Accugait, AMTI^®^). Participants’ upper body motion was assessed using a 3D motion analysis system (Codamotion^®^) including 4 infrared cameras. Each participant was equipped with 13 reflective markers fixed on specific anatomical landmarks (see Fig 1 for a detailed placement of markers). The same, trained, operator (orthopedic surgeon) positioned all markers for all patients and used the same following method to choose the markers’ position: T1 and L5 were easily located through simple palpation. T12-L1 (thoraco-lumbar junction), and the apex of the kyphosis and lordosis were determined by counting the vertebrae starting from T1. Recording systems were synchronized and sampled at 100 Hz. All data were low-pass filtered with a zero-lag fourth order Butterworth filter with a 10 Hz cut-off frequency.

**Fig. 1.**
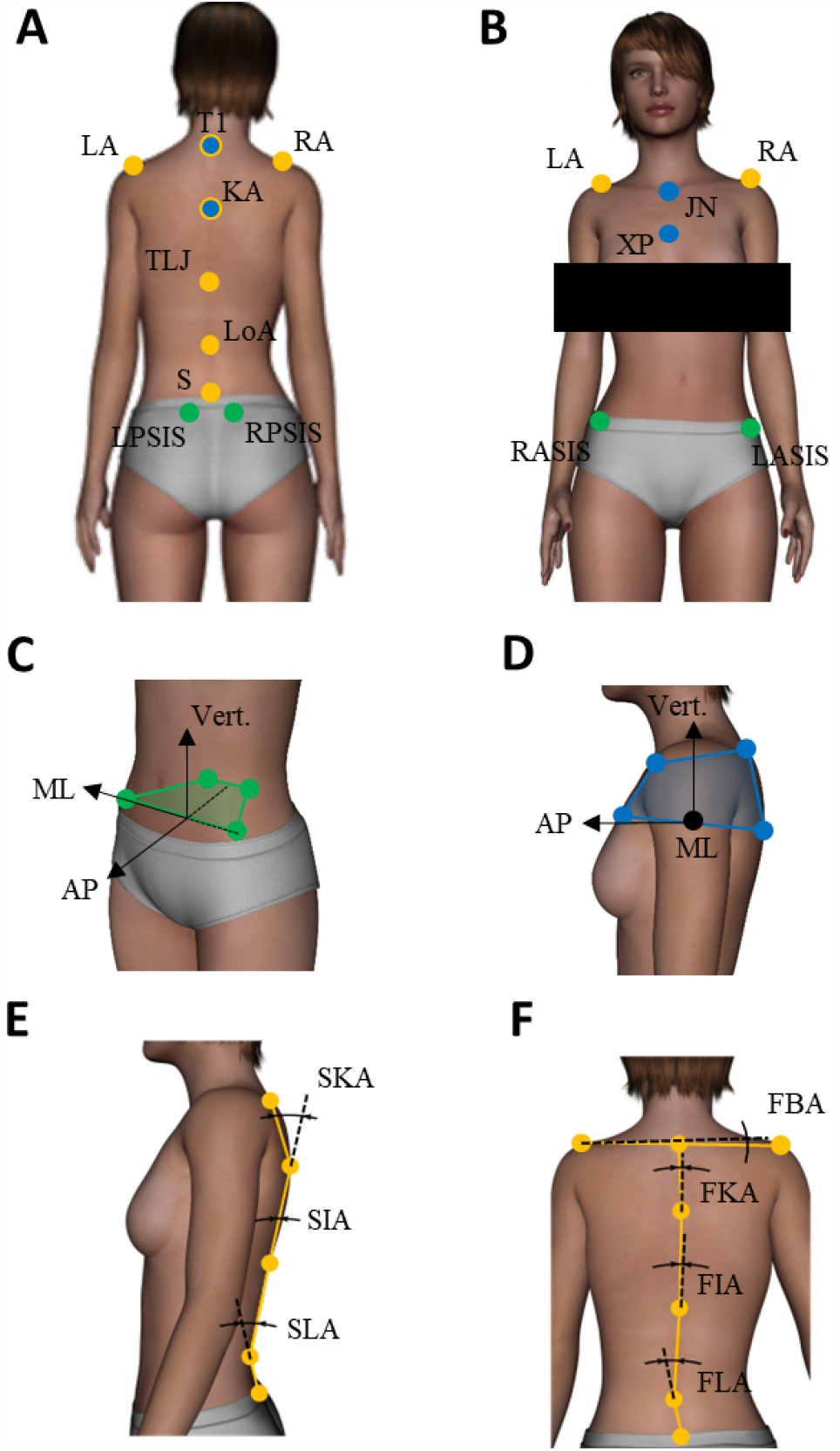
Reflective marker positions and dependent variables definition. (A + B) Back (A) and front (B) views of markers location. Reference frame was defined by its antero-posterior axis (AP), the horizontal symmetry axis between both foot, its medio-lateral axis (ML) perpendicular to AP axis in the horizontal plane, and its vertical axis, normal to the horizontal plane pointing upwards. (C) Definition of pelvic rigid body: ML axis is defined as the axis connecting RASIS and LASIS, vertical axis is the normal to the plane containing RASIS, LASIS and the middle of RPSIS and LPSIS then AP axis is the vector product between ML and vertical axis. (D) Definition of thoracic rigid body: vertical axis is defined as the axis connecting the middle of XP and KP and the middle of T1 and JN, ML axis is the normal to the plane containing T1, JN and the middle between XP and KM, then AP axis is the vector product between ML and vertical axis. (E) Definition of the sagittal angles of the spine. (F) Definition of the frontal angles of the spine and of the biacromial line, connecting LA and RA. Note that transverse rotation of biacromial line, and global angle of the spine with respect to the vertical in sagittal and transverse planes, which have also been investigated are not represented here. *LA = left acromion, RA = right acromion, T1 = first thoracic spinous process, KA = kyphosis apex, TLJ = thoracolumbar junction, LoA = lordosis apex, S = sacrum, LPSIS = left posterior-superior iliac spine, RPSIS = right posterior-superior iliac spine, RASIS = right antero-superior iliac spine, LASIS = left antero-superior iliac spine, XP = xyphoïd process, JN = jugular notch, SKA = sagittal kyphosis angle, SIA = sagittal inflexion angle, SLA = sagittal lordosis angle, FKA = frontal kyphosis angle, FIA = frontal inflexion angle, FLA = frontal lordosis angle, FAA = frontal biacromial angle*.

### Data analysis

Computation of all parameters and statistical analyses were performed with Matlab^®^ software. In static condition, participants’ stability was evaluated through 95 % confidence ellipse area, mean speed of CoP and sample entropy assessing the amount of attention invested in postural control [12]. Complying with optimization criterion, we chose m = 3 and r = 0.04 in the entropy algorithm. Pelvis and thoracic rigid bodies orientation, defined by Euler angles following ISB recommendations [13] – with respect to the reference frame – were investigated. To extend this evaluation and assess relevant spine morphology, different spine and acromial line angles were also computed [14] (see Fig. 1 and legend for precise definitions). Note that the anatomical landmarks on the spine were chosen relatively to normal kyphosis and lordosis and not to scoliotic curvature, so as to enable direct comparison between HS and AIS. Regarding dynamic tasks, angular range of motion (ROM), displacement of the pelvis and the thorax rigid bodies as well as ROM of spine angles between initial and final position of the subject were investigated in the plane in which motion was performed (frontal for lateral bending, sagittal for forward bending and transverse for rotations). AP and ML CoP displacements during the task were also investigated. Mean of each dependent variable was considered. All variables were expressed as mean ± standard error (SE) after verification of normal distribution and equal variance with Shapiro-Wilk test. To analyze presumed differences for the static and forward bending tasks, 3 groups (HS, AIS-LL, AIS-RT) ANOVAs were applied to the dependent variables. For lateral bending and rotation movement tasks, 3 groups (HS, AIS-LL, AIS-RT) x 2 sides (left, right) ANOVAs were performed.

## Results

Results are summarized in table 1.

There was no significant difference between groups in terms of age, height, and weight. AIS-LL and AIS-RT groups exhibited similar Cobb angles and skeletal maturity based on Risser sign.

Regarding static task, analysis of 95 % confidence ellipse area showed a main effect of group. Sway area was significantly larger for AIS-RT than for HS group, the AIS-LL group exhibiting an intermediary behavior. Mean speed of CoP displacements and sample entropy showed no significant effect between groups. Analysis of static segmental position of the upper body with respect to the reference frame showed a main effect of group for pelvis axial rotation (transverse angle). Amplitude of pelvis rotation was larger for AIS-RT than for AIS-LL group. HS group was not different from both AIS groups. A second main effect of group was present for sagittal kyphosis angle. It was significantly lower for AIS-RT group than for both HS and AIS-LL groups. Finally, although no significant effect of group for bi-acromial line rotation relatively to the pelvis was observed, around one third of all AIS patients showed a rotation of the bi-acromial line relatively to the pelvis greater than 5 deg. When present, this trunk rotation was always toward the left.

In lateral bending, analysis of kinematic data (Fig. 2) showed a significant main effect of group for pelvis ROM. It was lower for both AIS groups than for HS group. In addition, ML amplitude of CoP displacements also showed a significant effect of group with a ML amplitude higher for HS than for both AIS groups. A significant effect of group was also present for frontal spine angle, which was lower for AIS-RT group than for HS and AIS-LL groups. Finally, pelvis and thorax frontal displacement both revealed a main effect of group. Post-hoc tests showed that contralateral pelvis displacement was significantly larger for AIS-LL than for HS, and that thorax displacement was significantly lower for AIS-RT than for HS group. Despite small differences between AIS-LL and AIS-RT groups, reported differences in lateral bending reflected a different bending strategy present in both AIS groups in comparison to HS group. While HS rotated their spine mainly around the pelvis, AIS subjects performed the bending so that fixed rotation point during bending was localized approximately at L2 (Fig. 2). Neither side nor interaction effects were significant.

**Fig. 2:**
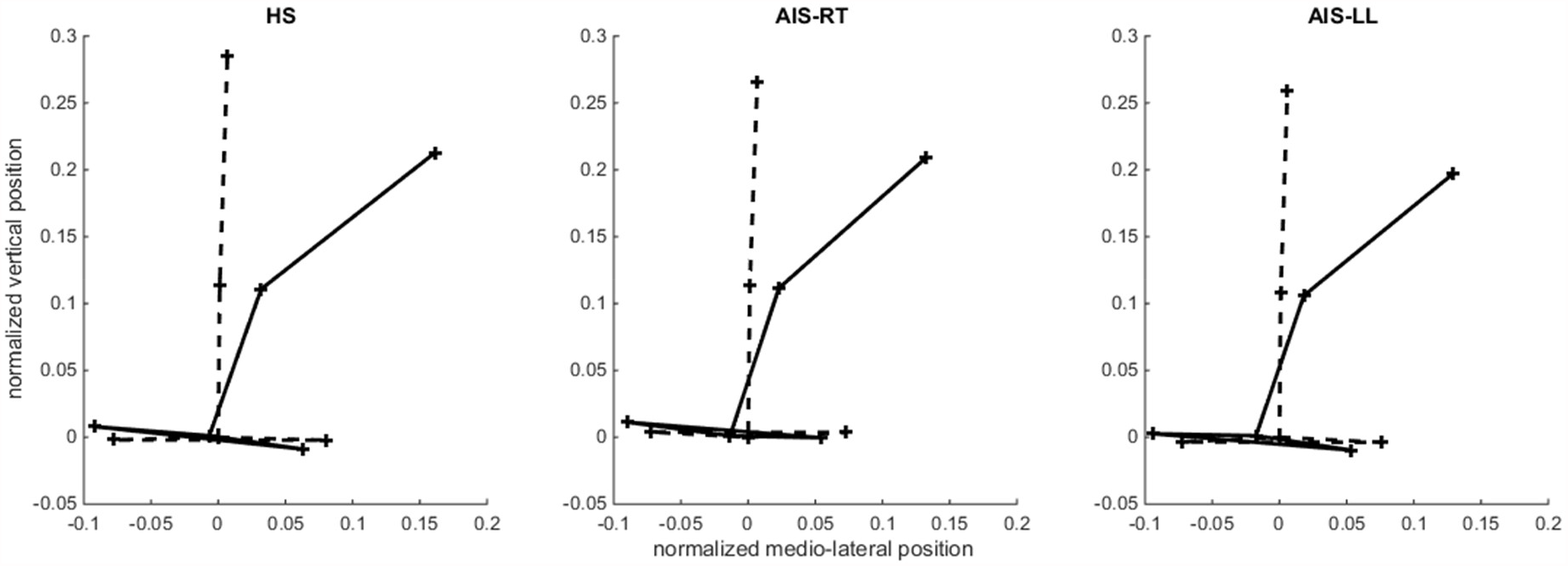
Back view of the spine in lateral bending. Plot of the back view (frontal) of the mean normalized positions of the spine (sacrum, inflexion point and T1) and the pelvis (antero-superior iliac spines) at the beginning (dotted line) and at the end (solid line) of the lateral bending for HS, AIS-LL and AIS-RT groups.

No significant differences were observed in rotation. However, ROM and displacements were consistently greater for HS than for AIS patients. In forward bending tasks, no differences were noted.

## Discussion

The first aim of this study was to extend postural differences traditionally observed in moderate to severe AIS patients to mild AIS patients. We showed that AIS-RT patients exhibited a greater sway area than HS, confirming previous results [4,5], while AIS-LL exhibited an intermediary behavior. CoP speed and sample entropy did not show any difference, suggesting that for the three groups postural control required a similar amount of muscular activity and attention. Regarding upper body kinematics during static task, we showed that mild scoliosis only sparsely affects body symmetry. However, when an asymmetry was observed, it was constantly in the same direction, with a trunk torsion toward the left with respect to the pelvis as already reported in previous studies for severe AIS-RT patients [2,3] and related to Cobb angle [8]. In our cohort, this trunk torsion – when present – was independent of AIS type and not linked to Cobb angle and could be considered as a predictive factor of scoliosis aggravation rather than severity. Results also highlighted a reduced kyphosis in AIS-RT with respect to healthy participants, that is, a decreased physiological sagittal curvature near the main scoliotic curvature – well known as the flat-back syndrome.

One of the main results concerned the motion differences between AIS patients and healthy subjects. AIS patients, whatever the type of scoliosis, showed a reduced pelvic rotation suggesting a reduced range of trunk motion in the frontal plane as reported during gait [7]. It could be due to an increased body stiffness because of 3D structural changes in the pelvis [15] and/or considered as a compensation for postural imbalance and/or to preserve patients from pain issues. Interestingly, lateral bending differences in pelvis and thorax displacements were indicative of different biomechanical strategies that seems similar in AIS-LL and AIS-RT. While bending, AIS performed a contralateral displacement of the pelvis and smaller displacement of the thorax so that fixed rotation point during bending was localized approximately at L2, whereas HS rotated mainly around the pelvis (Fig. 2). This strategy may be used to compensate for postural imbalance. Scoliotic patients would try limiting CoP displacements to avoid a potentially critical position as confirmed by the higher lateral amplitude of CoP displacements for healthy subjects. However, this strategy was not observable for frontal bending, certainly because it is a symmetrical movement less destabilizing.

Contrary to our hypotheses, no asymmetry in right and left motions were observed whether in AIS-LL or AIS-RT groups, suggesting a similar and symmetric kinematic behavior whatever the side of the curvature. The curve size of our patients may be also not sufficient to observe sub-group differences between curve types. Nevertheless, this indicated that the observed kinematic alterations are rather independent of static deformation, and already present at an early stage of AIS suggesting that they could be investigated as markers for early detection of AIS and/or aggravation. Finally, if kinematic analysis seems to be an efficient tool to characterize AIS, further investigations using multivariate methods on larger cohorts should be necessary to better characterize AIS motion and come up with a more satisfying three-dimensional classification of AIS, and potential markers for diagnosis.

## Data Availability

Data and analysis codes will be made available on request to the corresponding author.

## Acknowledgment

This work was supported by French state funds managed by the ANR within the Investissements d’Avenir programme (Labex CAMI) under reference ANR-11-LABX-0004, by a CNRS grant and by Chabloz Orthopédie.

## Data availability

Data and analysis codes will be made available on request to the corresponding author.

